# Health Perceptions of Adverse Weather in Older Adults in England: Analysis of 2019/20 Survey Data

**DOI:** 10.1101/2024.04.19.24306072

**Authors:** Grace Turner, Agostinho Moreira de Sousa, Emer O’Connell, Sari Kovats, Katya Brooks, Owen Landeg, Sharif Ismail, Anusha Rajamani, Shakoor Hajat

**Affiliations:** NIHR Health Protection Research Unit in Environmental Change and Health, London School of Hygiene and Tropical Medicine, UK; Extreme Events and Health Protection Team, UK Health Security Agency, UK

**Keywords:** Hot weather, cold weather, high/low temperatures, risk perception, behaviour, public health, older adults, England

## Abstract

**Introduction:** Health risks to vulnerable groups associated with hot and cold weather are well-documented. Older adults, aged 65 and above, are particularly vulnerable to higher and lower temperatures.

**Aim:** To explore older adult perception of health risks from high and low temperatures, what health-protective measures they have undertaken, and the factors associated with risks and responses.

**Methods:** Ipsos MORI conducted face-to-face surveys for the UK Health Security Agency with 461 participants in the cold weather survey and 452 in the hot weather survey. Participants reported temperature-related symptoms, risk perceptions for different groups, and behaviours during hot and cold weather. Data analysis involved using binomial logistic regression models to assess potential factors (demographics, vulnerability, behaviours, and responses) associated with older adults’ health risk perception in hot and cold weather.

**Results:** Less than half of older adults in both surveys agreed that hot or cold weather posed a risk to their health. Older adults with higher education, household income >£25,000 annually or home ownership were less likely to perceive their health at risk during cold weather. In both surveys, older adults who perceived people of the same age or people living alone as at an increased risk were more likely to perceive their own health as at risk. Furthermore, during cold weather, older adults were more likely to self-identify their health at risk when reporting other adults aged 65yrs+ to be at an increased risk, but not during hot weather. Various temperature-related protective behaviours were associated with older adults’ risk perception in hot and cold weather.

**Conclusion:** These findings highlight the need for effective risk communication strategies and targeted health messaging for older adults to support self-identification of risk. Future research should focus on barriers to risk perception and promoting health-protective behaviours in this population.

## Introduction

Risks to health from hot and cold weather are well documented(1, 2). Whilst evidence suggests a decrease in cold spells and frost days in the UK, the frequency and intensity of heatwaves is increasing(3). Risks to health from heat are identified as a key risk from climate change in the Third UK Climate Change Risk Assessment, published in 2021. However, the assessment also acknowledges that cold risks will remain significant to the end of the century(4–6). Climate change will increase the risk of extremely high outdoor temperatures, more consecutive hot days and heatwaves across regions in England (7, 8).

This paper focuses on the summer of 2019 and winter of 2020. The summer of 2019 saw 3 heatwaves with two Level 3 Heat Health Alerts (HHA) being issued, during the second heatwave (21st-28th July), an estimated 572 excess deaths were observed in 65+ year olds in England(9). In 2019-2020, approximately 3600 instances of overheating (>26°C) were reported by NHS Trust buildings in England(10). In 2020, an estimated 28,300 excess winter deaths including deaths from respiratory infections (excluding Covid-19) occurred in England and Wales in 2019/20 which was nearly 20% higher than 2018/19. In recent years, heatwaves in England have resulted in significant mortality (11). Recent analysis shows across England and Wales, from 2000 and 2019, an annual average of 800 heat-related deaths and 60,500 deaths associated with cold(12). Furthermore, in 2022, the UK experienced the hottest day on record (40.3°C), the first Level 4 HHA and there was an estimated 2,985 all-cause excess deaths associated with five heat episodes – the highest number in any given year(13).

Considering the social determinants of vulnerability to minimise temperature-related risks to health is vital, particularly amongst groups who may have difficulty adapting their environment or behaviours to the changing conditions, for example older adults, young children and those with long-term health conditions(6, 14). Older adults aged 65yrs+ have a higher risk of mortality and admission during periods of high and low temperatures so are the focus on the current study. The relative risk of death in older adults increases as temperatures reach extremes at both ends of the range: however, across a winter or summer season, due to the large number greater of days at moderate but harmful temperatures, the greatest health burden occurs outside of the extremes. The relative risk of mortality increased and negative health effects are observed at relatively moderate outdoor mean temperature change e.g. for cold this is seen between 4°C to 8°C (15, 16). Temperatures in excess of 25°C are associated with severe illness and excess heat-related deaths in vulnerable people(14). The health risks in older adults from changes in temperature are due to impaired thermoregulation, pre-existing health conditions and impaired ability to perform self-protective behaviours(17–20). Exposure to cold weather in older adults has been linked with increased risk of strokes, cardiovascular diseases and injuries e.g. falls. Furthermore, the key outcome of concern regarding heat exposure amongst older adults is the exacerbation of existing respiratory and cardiovascular conditions. The risks to health from hot and cold weather can be mitigated and action can be taken to minimise the impacts to human health through planning and emergency response strategies (21).

A scoping review of health risk perception in hot and cold weather amongst older adults concluded that older people often do not perceive themselves to be at an increased risk (3). Factors influencing personal health risk perception identified in the literature are: knowledge of hot and cold weather and associated health risks, presence of comorbidities, age and self-identity, perceived weather severity, previous experiences of adverse weather, perceived effectiveness of protective behaviours, and external locus of control (3). The evaluation of the Heatwave Plan (HWP) for England reported only 40% of 142 older adults aged 75+ years and 33% of 380 adults aged 65-74 year olds living in England agreed that hot weather was a risk to their health(22). Qualitative research suggests that while older people do not consider themselves “vulnerable” to heat or cold, they might perceive other older persons as vulnerable(23). A survey in England reported most older adults do undertake some heat protection measures that they consider “common sense” during heatwaves(^23^). Generally, evidence reports of limited knowledge of heat exhaustion, heat stress and exacerbation of existing respiratory, renal and cardiovascular diseases. Older adults and other vulnerable groups compared to the general population have demonstrated variable uptake of recommended behaviours e.g. drinking fluids, and opening or closing windows) during hot weather, staying active and dressing warmly in cold weather (18, 24, 25).

This paper aims to explore a representative sample of older adult’s (aged 65 years and over) perception of the risks from both high and low temperatures, what health-protective measures they have undertaken, and the factors associated with risks and responses. This paper is the first analysis of both hot and cold weather risk perception and compares the potential synergies and differences in factors such as region, age, income, influencing older adults’ risk perception of their health in England. Furthermore, this study is the first to determine links between adopting temperature-related behaviours and predicting personal health risk perception in older adults. The sample population is representative of England which supports a detailed analysis of the development of insight-informed action. This is useful for national planning and focusing hot and cold weather messaging around behaviour change techniques, prioritising communication of actions to take and how behaviour interacts with health perception. This analysis also explores older adult perceptions of other older adults and vulnerable groups which is useful to understand and inform how public health messaging should target older adults as a vulnerable group e.g. terminology during extreme temperatures change. Within the research priorities of the Temperature chapter in the Health Effects of Climate Change report published by the UK Health Security Agency (UKHSA), further analysis is required to understand the risks from hot and cold weather on vulnerable groups including older adults(6). Results and recommendations from this paper will crucially feed into the evidence and messaging updates for the published UKHSA Adverse Weather and Health Plan which was launched in April 2023(26).

## Materials and Methods

### 1.1. Survey and study participants

We analysed data collected from two surveys conducted by Ipsos MORI that affect heat and cold health risk perception in older adults (aged 65 years and older) in England, commissioned by UKHSA. Research objectives were to:

1. Identify factors associated with adverse weather health risk perceptions
2. Identify responses or behaviours associated with hot/cold weather risk perception.

The heat survey (n = 1706) was conducted from 26th July to 4th August 2019 following very hot weather in the UK and Europe and the cold survey (n = 1719) took place between 7th–23rd February 2020 following a mild winter. The survey was specifically commissioned to explore the public’s perceptions, awareness and experience of the risks of hot weather in England. Both surveys were conducted in person by an Ipsos researcher in English using a nationally representative sample of the population aged 15 and over in England. Quotas were set for age by gender, region, working status and housing tenure. This paper presents the results only in older adults aged 65 years and over.

Participants were asked questions relating to their perceptions, awareness and experience of the risks of recent hot or cold weather events in 2019/20. Initially in both surveys, participants personal risk perception was captured by asking them to respond on a Likert scale to what extent they agreed with the following statement “hot/cold weather is a risk to my health”. The surveys cover three themes: participant temperature-related symptoms; participant health risk perception of other groups (e.g. the very young); and acute participant responses or behaviours to hot and cold weather event (Supplementary material S1). Questions were a mixture of closed yes/no, Likert scale and listed options for selection. Temperature-related symptom outcomes were not included in the regression analysis but are presented in the descriptive data.

### 1.2. Analysis

IBM SPSS [Version 28.0.0.0] was used to re-code and categorise data. The analysis was conducted in R [Version 1.3.1093]. First, descriptive and exploratory analyses were carried out (cross tabulations and bivariate correlation matrix) to identify variable relationships. Models were selected from running ordinary least squares regression models (OLSR) with all potentially relevant predictor variables to identify which independent variables best predict older adults’ perception of risk to their health from hot/cold weather. OLSR models which reported the lowest Akaike Information Criterion (AIC) value and highest adjusted R-squared value were used in the final logistic regression analyses.

*Binomial logistic regression models* were fit to assess potential factors associated with older adults health risk perception in hot or cold weather. The dependent variable for each analysis was comparing agreement (strongly agree/agree) that “hot/cold weather is a risk to my health” (1) versus disagreeing (strongly disagree/agree) or being neutral (neither agree nor disagree) in response to “hot/cold weather is a risk to my health” (0). The full list of independent variables is in Supplementary material S2.

Three analyses were carried out:

- Analysis 1: Demographic determinants of older adult health risk perception of hot/cold weather
- Analysis 2: Older adult’s perception of other groups health risk in hot/cold weather as a predictor of own personal risk
- Analysis 3: Older adult responses or adopted behaviours association with their health risk perception in hot/cold weather

Each analysis was conducted for each survey. Each analysis adjusted for the following covariates: age, sex, education, annual income, home ownership and region (South vs Midlands/North). Ethnicity was not included as a covariate due to a small sample size of non-white British.

### 1.3. Ethics

The data collection process was carried out by Ipsos MORI, a market research company, who provided the following statement: “This project was reviewed by the internal IPSOS MORI Ethics Group, but it was not submitted externally for ethics approval which is standard practice in market research where the research topic is not considered sensitive and the participants are not considered vulnerable or unable to give informed consent. It was also judged that there would be no potential disclosure of harm through the survey. The data received by the authors in this study was anonymised and individuals are not identifiable”. Recruitment for each study took place during the following time windows: heat survey 26th July to 4th August 2019; cold survey 7th - 23rd February 2020. Verbal consent was sought and received from each participant at the time of the survey and documented by the Ipsos MORI research on the participant response sheet for each.

## Results

Table 1 describes the characteristics of the sub-sample of older adults in each survey (heat survey n = 452 (26%), cold survey n = 461 (27%). Just under half of older adults in both surveys agreed with the statement “hot/cold weather is a risk to my health” (47% heat survey, 45% cold survey).

**Table 1:** Demographic information of survey participants. Older adults (Aged 65 years and over).

| Variable | Heat survey<br>participants, n (%) | Cold survey<br>participants, n (%) | General pop.<br>Distribution |
| --- | --- | --- | --- |
| All survey participants (n) | 1706 | 1719 | - |
| **Older adults (n)** | 452 (26%) | 461 (27%) | 18.3% |
| **Age** | Mean = 74 | Mean = 74 | Mean = 73 |
| 65-74 | 241 (14%) | 274 (16%) | 9.8% |
| 75-84 | 170 (9%) | 135 (7%) | 6.1% |
| 85+ | 41 (2%) | 52 (3%) | 2.4% |
| **Sex:** |  |  |  |
| Male | 219 (48%) | 257 (56%) | 46% |
| Female | 233 (52%) | 204 (44%) | 54% |
| **Education:** |  |  |  |
| No formal education | 107 (24%) | 91 (20%) | 18.2% |
| GCSE/O-level/A-level/Vocational<br>qualification | 173 (38%) | 188 (40%) | 30.3% |
| Higher education (Degree or above) | 121 (27%) | 146 (31%) | 33.8% |
| Other | 51 (11%) | 36 (8%) | 8.1% |
| **Annual income:** |  |  |  |
| 0 - £24,999 | 146 (32%) | 184 (40%) | - |
| £25,000 - £49,999 | 85 (19%) | 87 (19%) | - |
| £50,000 + | 38 (8%) | 33 (7%) | - |
| Unknown or refused | 183 (41%) | 157 (34%) | - |
| **Ethnicity:** |  |  |  |
| White British | 423 (94%) | 440 (95%) | 90% |
| Non-White British | 29 (6%) | 19 (5%) | 10% |
| **Employment status:** |  |  |  |
| Employed | 43 (9%) | 41 (8%) | 10% |
| Retired | 409 (91%) | 420 (91%) | 90% |
| **Home ownership:** |  |  |  |
| Home owned accommodation | 380 (84%) | 368 (80%) | 77% |
| Rented accommodation | 72 (16%) | 85 (20%) | 23% |
| **Region:** |  |  |  |
| North East | 21 (5%) | 22 (5%) | 5% |
| North West | 68 (15%) | 61 (13%) | 13% |
| Yorkshire & the Humber | 47 (10%) | 39 (8%) | 10% |
| East Midlands | 38 (8%) | 52 (11%) | 9% |
| West Midlands | 41 (9%) | 39 (8%) | 11% |
| East | 68 (15%) | 59 (13%) | 12% |
| London | 62 (14%) | 44 (10%) | 10% |
| South East | 61 (14%) | 68 (15%) | 17% |
| South West | 46 (10%) | 77 (17%) | 12% |

### Heat and cold-related symptoms

In both surveys, older adults reported experiencing at least one symptom during hot (76%) or cold weather (40%). The most commonly reported heat-related symptoms included experiencing difficulty keeping cool and feeling too hot (40%), having difficulty sleeping (52%) and physical symptoms such as a headache, dehydration or sunburn (11%). Older adults reported fewer cold-related symptoms (60% reported no symptoms following 2019 winter).

#### Analysis 1: Demographic determinants of older adult health risk perception in hot/cold weather

Regional differences in older adult health risk perception dependent on hot or cold weather across England are observed between the surveys (Supplementary material S3). Across the Midlands, East of England and Yorkshire & Humber, a higher percentage of respondents perceived their health to be at risk in hot weather than cold weather, ranging from 47% - 63% compared to 29% - 52% respectively. Little to no difference in older adults health risk perception in the North East, South East, South West or London in either the hot weather or cold weather survey responses. In the North West, 64% of older adults in the cold weather survey agreed their health was at risk during cold weather compared to 46% of older adults in the hot weather survey.

Older adults who had a degree or higher (OR 0.71, 95% CI [0.56-0.89]), household income over £25,000 a year (OR 0.75, 95% CI [0.60-0.94]) and owned their own home (OR 0.64, 95% CI [0.51-0.79]) were significantly less likely to perceive their health as at risk in cold weather (Figure 1). Older adults who lived in the South of England (South West, South East or London) were 19% less likely to perceive their health to be at risk during hot weather compared to those living in the Midlands or Northern England (OR 0.81, 95% CI [0.66-0.99], however the association is weak (p = 0.043). No other determinant (such as age or income) predicted older adults perceiving themselves at risk from high temperatures.

**Figure 1:**
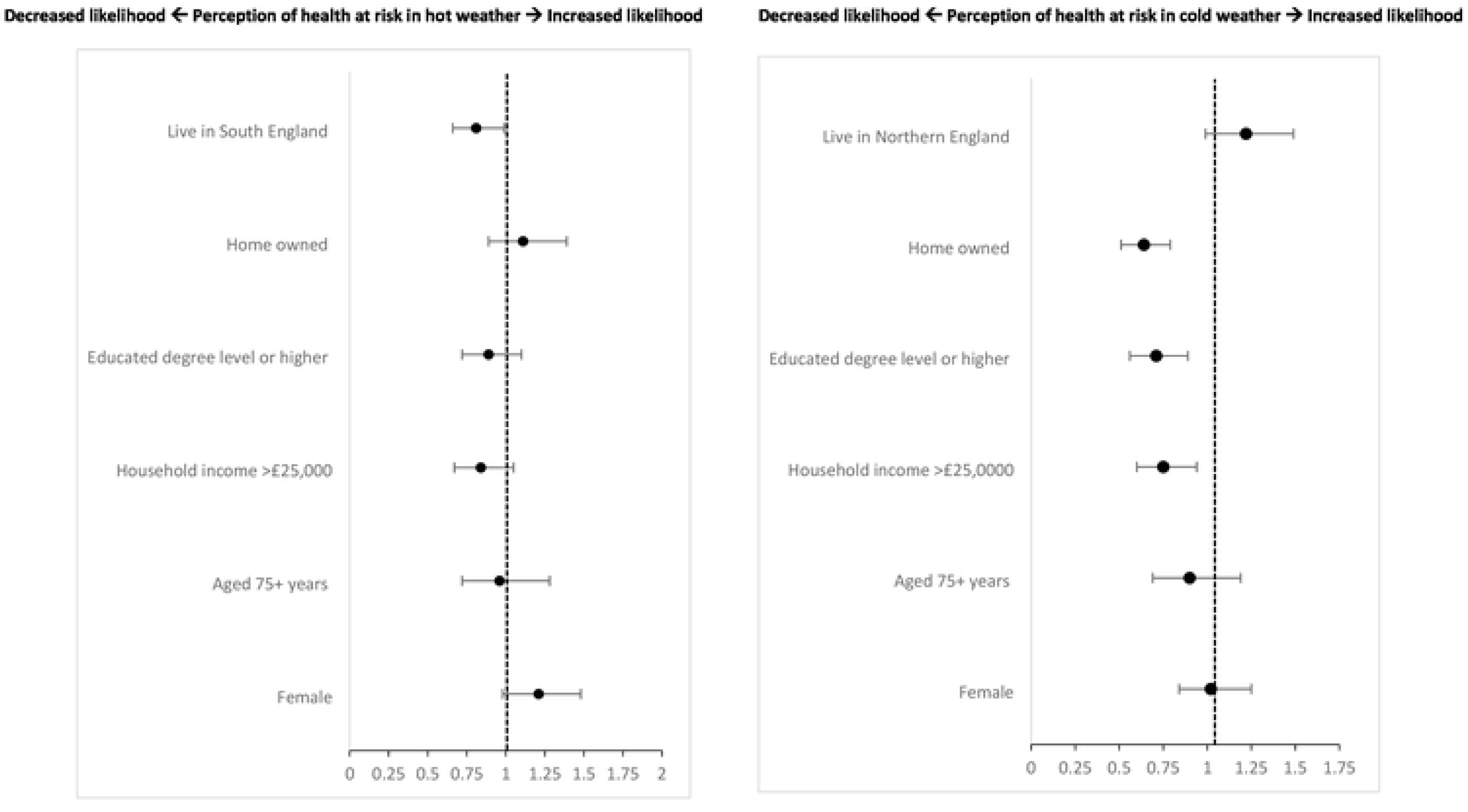
Analysis 1 – Binomial logistic regression model plot showing demographic predictors of older adult (aged 65+ years) perception of risk to health from hot (left) and cold (right) weather. Figure shows odds ratios. * = p <0.05, ** = p<0.001. Adjusted for age, sex, education, income, home tenancy and region.

#### Analysis 2: Older adult’s perception of other vulnerable groups health risk in hot/cold weather

A higher percentage of older adults who perceived their own health as at risk in hot weather also identified different vulnerable groups as at an increased risk (Supplementary material S2). In both surveys, a larger proportion of older adults who disagreed that their own health is at risk also disagreed that ‘people aged 65 years and over’ were at risk, however, less disagreed with ‘people of the same age’ being at risk. It appears that there is varying consistency regarding older adults’ awareness of their own health risk and risk perception of other vulnerable groups in relation to high and low temperatures.

Figure 2 presents the association between older adults’ perception of other vulnerable groups health risk as predictors of personal risk perception in high/low temperatures. In both survey results, older adults that identified people of the same age (Heat: OR 3.75, 95% CI [2.85-4.95]; Cold: OR 4.17, 95% [3.12-5.58]) and people living alone (Heat: OR 1.94, 95% CI [1.45-2.59]; Cold: OR 1.41, 95% CI [1.08-1.84]) as at an increased risk were significantly more likely to perceive their own health as at risk. Additionally, older adults perceiving others aged 65+ as at risk in cold weather was significantly associated with older adults perceiving their own health to be at risk (OR 1.4, 95% CI [1.06-1.84]). Identifying babies and infants (OR 1.48, 95% CI [1.16 – 1.87]) as at an increased risk in cold weather was a significant predictor of older adults perceiving their own health as at risk.

**Figure 2:**
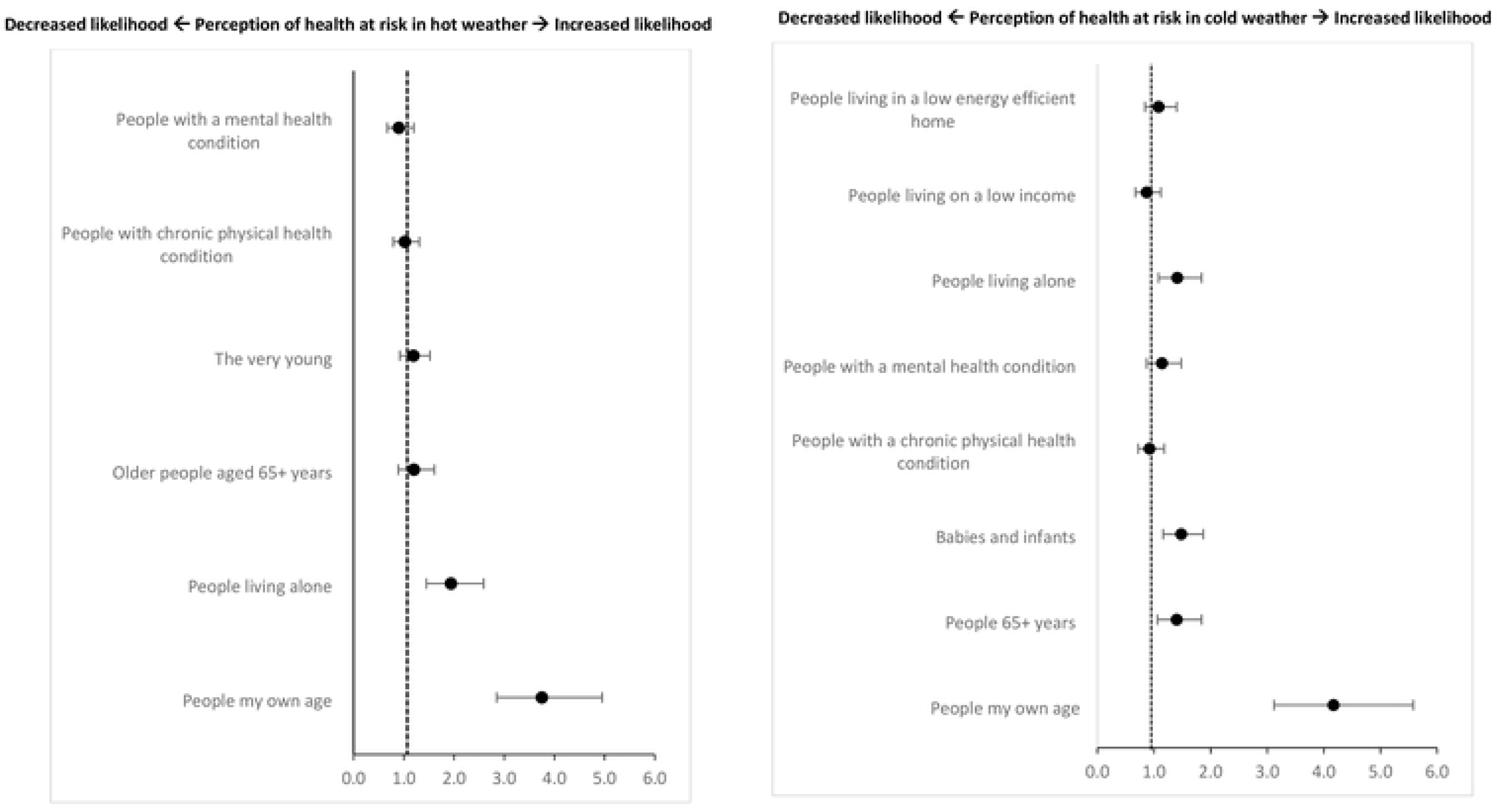
Analysis 2 – Binomial logistic regression model plot showing perception of other vulnerable groups risk in hot/cold weather as predictors of older adults (aged 65+years) perception of risk to health from hot (left) and cold (right) weather. Figures show odds ratios. * = p < 0.05, ** = p <0.001. Adjusted for age, sex, education, income, home tenancy and region.

#### Analysis 3: Older adult responses or adopted behaviours associated with their health risk perception in hot/cold weather

Across surveys, there were no notable differences in the proportion of adults reporting at least one action to reduce the impact of cold (89%) and the impact of heat (90%). More than half of older adults reported the following heat-related behaviours: drinking more fluids (84%), opening windows at night/cooler parts of the day (61%), wearing loose clothing and/or a hat (59%), finding somewhere that felt cool (53%) and keeping curtains closed on windows exposure to direct sunlight (50%). Over half of older adults reported the following cold-related responses: layering clothing (80%), heating home >18°C (77%), having their boiler checked by an engineer (61%), heating rooms most occupied (59%), keeping bedroom window closed at night (59%), checking the forecast and planning ahead (53%) and drinking warm drinks (58%). All other behaviours carried out during hot or cold weather were reported by <10% of respondents in either survey.

We investigated whether health-protective responses and behaviours to hot and cold weather were associated with older adults’ perception of risk (Figure 3). Older adults who reported using or buying a fan during hot weather were 43% more likely to agree their health to be at risk in hot weather (OR 1.43, 95% [1.11-1.84]). Older adults who reported staying indoors (OR 2.06, 95% CI [1.55-2.74]) or limited physical activity to cooler parts of the day (OR 1.48, 95% CI [1.14-1.92]) were significantly more likely to perceive their health as at risk in hot weather. Keeping curtains or windows closed that are exposed to direct sunlight were statistically significant predictors for older adults being more likely to agree their health is at risk in hot weather (OR 1.37, 95% CI[0.31 – 1.77]). In cold weather, older adults who stocked up on food and medicine (OR 2.54, 95% CI [1.98-3.27]) were more likely to perceive their health to be at risk. Finally, older adults who checked the forecast and planned ahead were 23% less likely to agree that their health is at risk during cold weather (95% CI [0.62-0.95]).

**Figure 3:**
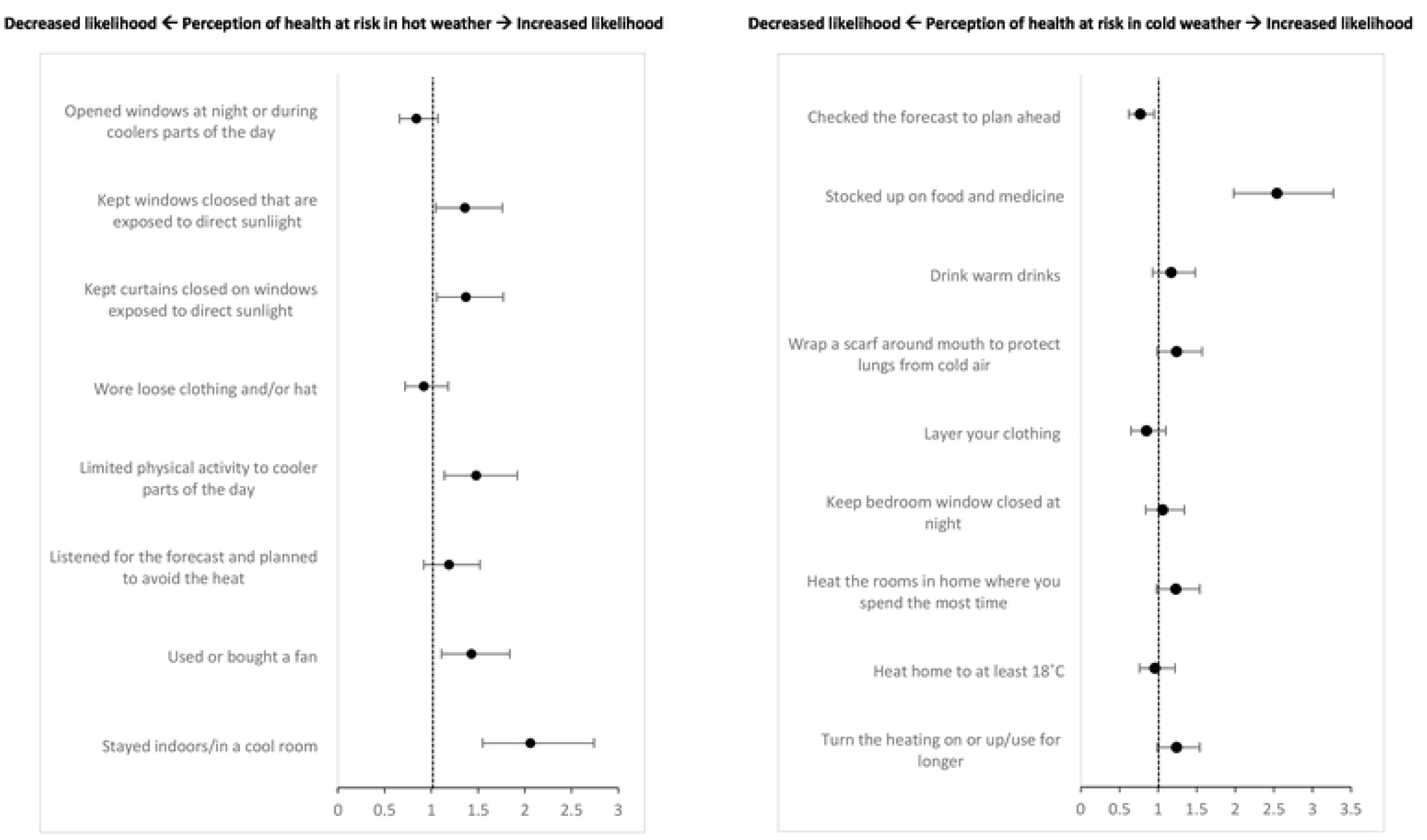
Analysis 3 – Binomial logistic regression model plot showing temperature-related behaviours as predictors of or older adults (aged 65+years) perception of risk to health from hot (left) and cold (right) weather. Figures show odds ratios. * = p < 0.05, ** = p <0.001. Adjusted for age, sex, education, income, home tenancy and region.

## Discussion

Our findings align with previous evidence that less than half of older adults in England consider their health to be at an increased risk in hot or cold weather events(3, 27, 28). In the evaluation of the HWP, fewer older adults reported their health as at risk in hot weather compared to this more recent survey. This finding may be due to the increased frequency and intensity of hot weather events in recent years (22). Low health risk perception in older adults could be attributed to individual conceptualisation of age-identity, past experiences, stoic attitudes, lack of understanding of health relevance or misconceptions of public health messaging(3, 29). Evidence in several Western countries suggests lowered risk perception due to a desire to embrace hot weather – typical in British culture (25, 30, 31). Although, in places such as Hong Kong, there is suggestive evidence of older adults being increasingly aware of the risks associated with their health from heat(29). Determining whether an increase in heatwaves has led to a rise in older adults health risk awareness may be useful to establish the relationship between the likelihood of experiencing extreme weather events more frequently and intensively and existing cultural norms in undermining individual action and response.

In this study, several demographic factors were associated with older adult health risk perception in hot or cold weather. Older adults with higher education, income, or home ownership were less likely to perceive their health at risk during cold weather. This may be due to fewer barriers and more resources for behaviour adaptation and mitigation of cold impacts for example being able to afford to heat home for longer or living in higher quality housing. Lower earners and less educated individuals were less aware of temperature-related behaviour change and had limited resources for health protection during adverse weather which aligns with previous research(18, 32). In the hot weather survey, older adults in the South of England were 19% less likely to perceive themselves at risk compared to those in the Midlands or North. Regional differences in health risk perception were observed, possibly due to behavioural differences during peak sun hours or fewer experiences of heat events in the Midlands and North of England. Existing literature shows there to be a north south divide in projected heat related deaths in England(33), although another study reported greater mortality associated with cold in northern and western regions(12). A previous study showed residents in the North of the UK were less likely to avoid the sun or walk in the shade(34). Older adults in the South of England may be more familiar with modifying their behaviour during hot weather, influencing their perception of health risks. Gender and age differences in risk perception were not observed in this analysis, contradicting previous research(18) that showed women above 75+ were more likely to recognize the risk of hot weather to their health, with no significant differences in the 65-74 age bracket(22).

In both surveys, older adults who identified people of the same age at increased risk were 3 times more likely to perceive themselves as at risk during hot or cold weather. However, when asked if older people aged 65+ were at risk, this did not predict personal risk perception in hot weather but did in cold weather. This suggests that older adults are more likely to respond to messages that represent people who they identify as similar to themselves but less with messaging describing the characteristics of vulnerability i.e. aged 65+ despite being in the same age category. This lack of consideration for the relationship between advanced aging and heightened risk in adverse weather is observed among some older adults, however, the finding in this paper highlights the need to explore the relationship between self-identify and characteristics of vulnerability further (35, 36). Heat and cold weather health messaging that targets older adults based on age as a vulnerability factor may hinder behaviour change due to misalignment with individual age identity. Identifying people living alone as at increased risk was associated with older adults perceiving their own health as at risk in both hot and cold weather surveys. Older adults who live alone are more likely to experience social isolation and may have limited access to support or healthcare services, which can heighten their perception of health risks during adverse weather(37) (38). There is evidence suggesting a possible disparity in older adults’ understanding of how comorbidities can worsen their health risk in extreme temperatures and whether they take action and follow recommended guidance(27, 31, 39, 40). It is important to identify the most vulnerable individuals and provide adequate care services to minimize the adverse impacts of hot and cold temperatures among older adults, especially those who may be isolated, in poor health, and lacking support.

In both surveys, variation in the uptake of key recommended acute health-protective behaviours or measures by older adults was observed e.g. limited reporting of regularly applying SPF or avoiding alcohol during hot weather, supported by previous evidence(24). However, the most common heat-related behaviours in this study (e.g. drinking cool fluids, opening windows at night) vary in reported uptake in previous research, many behaviours were poorly adopted by older adults outlined in the Heatwave Plan for England (3, 24, 25). This variation in reported behaviour during hot weather could indicate that older adults adopt key heat adaptive behaviours however, this is not always captured in research as responses are dependent on the survey question and context i.e. older adult’s motivation for carrying out certain behaviours may not be related to health protection and therefore fail to report. Furthermore, some behaviours may not be successfully adopted due to social barriers e.g. UK alcohol drinking culture. A survey of potentially vulnerable groups reported that avoiding alcohol (amongst other actions) was not perceived as an effective heat protection behaviour(25).

In this study, heat-related behaviours associated with older adults perceiving increased health risk in high temperatures included changing activity and modifying the home environment. Previous research has shown that these behaviours are often motivated by alleviating discomfort from heat exposure and reducing UV exposure(25, 27, 41, 42). Behaviours associated with an increase in risk perception in hot weather were primarily related to home modifications such as closing windows, curtains, using fans, and creating a cool room. In contrast, altering indoor temperature and closing windows were not associated with risk perception in the cold survey. This difference may be due to the routine behaviour of warming the home during cold weather compared to conscious responses to cool home during hot weather to alleviate thermal discomfort. Checking the forecast to plan for cold weather was associated with a reduced odds of perceiving health risk, likely due to reasons unrelated to health, such as daily planning. Older adults may view certain acute health protection behaviours as common sense, dismissing the severity of potential health risks as their motivation for adopting these behaviours. Lastly, older adults who actively stocked up on food and medicine due to cold weather were 2.5 times more likely to perceive their health as at risk. This suggests that those who engage in health-protective behaviours are likely to be more health conscious and may be older adults who live alone or have poor health(22). It is important to note that there are many other effective mitigations e.g. greening walls, insulating homes however, these are longer term, strategic prevention messages. The findings from this study are important for acute public health messaging during the immediate onset of a heat or cold event, as this is a shorter window (e.g. 48 hours prior to event) for communicating action which therefore focuses on immediate actions older adults can take to protect their health.

### Strengths and policy implications

This is a comprehensive study of health risk perception in hot and cold weather amongst older adults in England. The survey was conducted in person, In English and was nationally representative. The results from this study show that perception of the risks of hot and cold weather remain low, and protective actions are varied, even among high risk populations such as older adults. Furthermore, the awareness of public messaging about the risks of hot weather are low. The findings from this analysis are important for public health agencies for implementing successful risk communication strategies, national, minister led public health campaigns as well as guiding future research as called for following the Environmental Audit Committee inquiry published in early 2024 (See Box 1) (43). Additionally, including survey data from both a hot weather and cold weather event allowed for a comparison of the differences in older adult risk perception in differing temperatures and the predicting factors which may have influence.

### Limitations

A limitation is that respondents’ health status was not captured in the survey, so we couldn’t compare risk perception between healthier and less healthy older adults. However, we did explore age ranges (65-74 vs 75+) as a proxy for frailty and found no significant difference in risk perception for hot/cold weather. July 2019 had three heatwaves and above-average rainfall in England, which may have affected participants’ recollection of heat-related behaviours. The cold weather survey followed a mild winter, possibly explaining low reporting of behaviours. The sample of older adults was not representative for ethnicity, with over 96% being White British. Ethnicity has been cited in the evidence as a possible risk modifier; however, it was not possible to assess this aspect due to the small proportion of non-white British people in the sample. Urban/rural information was also not captured, which is important for understanding health risk perception in cold weather (e.g., healthcare access disruptions from snow or ice). It would be useful to investigate if living in rural areas affects older adults’ temperature-related health risk perception.

### Conclusion

This study found that most older adults do not perceive their health as at risk in hot or cold weather. The analysis identified demographic predictors of health risk perception in older adults for both hot (region) and cold weather (income, education, home ownership). Differences were observed in how older adults view other vulnerable groups at risk in hot and cold weather, depending on the language used to describe older people and how public health messaging targets vulnerable groups. Both surveys showed variation in the uptake of health-protective behaviours among older adults and some health-protective behaviours reported by older adults were associated with perceiving their health as at an increased or decreased risk in hot or cold weather. Persistent barriers exist for older adults in recognising their increased health risk during hot and cold weather. These barriers may be due to self-identification of vulnerability, awareness of health risks, stoicism, cultural norms, language/terminology used in messaging, and demographic and regional factors. These findings are important for public health agencies to implement effective risk communication strategies and health messaging for older adults (box 1). Future research should further examine barriers to risk perception and adoption of health-protective behaviours in older adults.

## Data availability

The data underlying the results presented in the study are available from (https://www.ipsos.com/en).

## References

1. Campbell S, Remenyi TA, White CJ, Johnston FH. Heatwave and health impact research: A global review. Health Place. 2018;53:210–8.

2. Ryti NR, Guo Y, Jaakkola JJ. Global Association of Cold Spells and Adverse Health Effects: A Systematic Review and Meta-Analysis. Environ Health Perspect. 2016;124(1):12–22.

3. Ratwatte P, Wehling H, Kovats S, Landeg O, Weston D. Factors associated with older adults’ perception of health risks of hot and cold weather event exposure: A scoping review. Frontiers in Public Health. 2022;10.

4. Curtis S, Fair A, Wistow J, Val DV, Oven K. Impact of extreme weather events and climate change for health and social care systems. Environmental Health. 2017;16(1):128.

5. UK Climate Risk. High Temperatures Briefing Findings from the third UK Climate Change Risk Assessment (CCRA3) Evidence Report 2021. London, UK; 2021.

6. Macintyre HL, Murage P. Health Effects of Climate Change (HECC) in the UK: 2023 report Chapter 2. Temperature effects on mortality in a changing climate London, UK; 2023.

7. Sahani J, Kumar P, Debele S, Emmanuel R. Heat risk of mortality in two different regions of the United Kingdom. Sustainable Cities and Society. 2022;80:103758.

8. Thompson R, Landeg O, Kar-Purkayastha I, Hajat S, Kovats S, O’Connell E. Heatwave Mortality in Summer 2020 in England: An Observational Study. Int J Environ Res Public Health. 2022;19(10).

9. PHE. PHE heatwave mortality monitoring Summer 2019 London, UK; 2019.

10. Brooks K, Landeg O, Kovats S, Sewell M, OConnell E. Heatwaves, hospitals and health system resilience in England: a qualitative assessment of frontline perspectives from the hot summer of 2019. BMJ Open. 2023;13(3):e068298.

11. Guo Y, Gasparrini A, Li S, Sera F, Vicedo-Cabrera AM, de Sousa Zanotti Stagliorio Coelho M, et al. Quantifying excess deaths related to heatwaves under climate change scenarios: A multicountry time series modelling study. PLOS Medicine. 2018;15(7):e1002629.

12. Gasparrini A, Masselot P, Scortichini M, Schneider R, Mistry MN, Sera F, et al. Small-area assessment of temperature-related mortality risks in England and Wales: a case time series analysis. Lancet Planet Health. 2022;6(7):e557–e64.

13. UKHSA. Heat mortality monitoring report: 2022. London, UK; 2023.

14. Berrang Ford L, Moreira Sousa A. Come rain or shine, adverse weather matters for our health London, UK; 2023.

15. Gasparrini A, Guo Y, Hashizume M, Lavigne E, Zanobetti A, Schwartz J, et al. Mortality risk attributable to high and low ambient temperature: a multicountry observational study. The Lancet. 2015;386(9991):369–75.

16. Chalabi Z, Erens, B., Hajat, S., Heffernan, C., Jones, L., Mays, N., Ritchie, B., Wilkinson, P.,. Evaluation of the implementation and health-related impacts of the Cold Weather Plan for England 2012 Final report. London, UK; 2015.

17. Millyard A, Layden JD, Pyne DB, Edwards AM, Bloxham SR. Impairments to Thermoregulation in the Elderly During Heat Exposure Events. Gerontol Geriatr Med. 2020;6:2333721420932432.

18. Khare S, Hajat S, Kovats S, Lefevre CE, de Bruin WB, Dessai S, et al. Heat protection behaviour in the UK: results of an online survey after the 2013 heatwave. BMC Public Health. 2015;15(1):878.

19. Nunes AR. General and specified vulnerability to extreme temperatures among older adults. International Journal of Environmental Health Research. 2020;30(5):515–32.

20. Kenny GP, Yardley J, Brown C, Sigal RJ, Jay O. Heat stress in older individuals and patients with common chronic diseases. Cmaj. 2010;182(10):1053–60.

21. PHE. The Cold Weather Plan for England. London, UK; 2015.

22. Williams L, Erens B, Ettelt S, Hajat S, Manacorda T, Mays N. Evaluation of the Heatwave Plan for England. London, UK; 2019.

23. Abrahamson V, Wolf J, Lorenzoni I, Fenn B, Kovats S, Wilkinson P, et al. Perceptions of heatwave risks to health: interview-based study of older people in London and Norwich, UK. J Public Health (Oxf). 2009;31(1):119–26.

24. PHE. Heatwave plan for England Protecting health and reducing harm from severe heat and heatwaves. London, UK; 2014.

25. Erens B, Williams L, Exley J, Ettelt S, Manacorda T, Hajat S, et al. Public attitudes to, and behaviours taken during, hot weather by vulnerable groups: results from a national survey in England. BMC Public Health. 2021;21(1):1631.

26. UKHSA. Adverse Weather and Health Plan Protecting health from weather related harm 202 to 2024. London, UK; 2023.

27. Abrahamson V, Wolf J, Lorenzoni I, Fenn B, Kovats S, Wilkinson P, et al. Perceptions of heatwave risks to health: interview-based study of older people in London and Norwich, UK. Journal of Public Health. 2008;31(1):119–26.

28. Howe PD, Marlon JR, Wang X, Leiserowitz A. Public perceptions of the health risks of extreme heat across US states, counties, and neighborhoods. Proceedings of the National Academy of Sciences. 2019;116(14):6743–8.

29. Lai ETC, Chau PH, Cheung K, Kwan M, Lau K, Woo J. Perception of extreme hot weather and the corresponding adaptations among older adults and service providers-A qualitative study in Hong Kong. Front Public Health. 2023;11:1056800.

30. Banwell C, Dixon J, Bambrick H, Edwards F, Kjellström T. Socio-cultural reflections on heat in Australia with implications for health and climate change adaptation. Glob Health Action. 2012;5.

31. Lane K, Wheeler K, Charles-Guzman K, Ahmed M, Blum M, Gregory K, et al. Extreme heat awareness and protective behaviors in New York City. J Urban Health. 2014;91(3):403–14.

32. Semenza JC, Hall DE, Wilson DJ, Bontempo BD, Sailor DJ, George LA. Public perception of climate change voluntary mitigation and barriers to behavior change. Am J Prev Med. 2008;35(5):479–87.

33. Jenkins K, Kennedy-Asser A, Andrews O, Lo YTE. Updated projections of UK heat-related mortality using policy-relevant global warming levels and socio-economic scenarios. Environmental Research Letters. 2022;17(11):114036.

34. McLoughlin N, Howarth C, Shreedhar G. Changing behavioral responses to heat risk in a warming world: How can communication approaches be improved? WIREs Climate Change. 2023;14(2):e819.

35. Wolf J, Adger WN, Lorenzoni I. Heat Waves and Cold Spells: An Analysis of Policy Response and Perceptions of Vulnerable Populations in the UK. Environment and Planning A: Economy and Space. 2010;42(11):2721–34.

36. Barak B, Schiffman LG. Cognitive Age: a Nonchronological Age Variable. Advances in Consumer Research. 1981;8(1):602–6.

37. Kafeety A, Henderson SB, Lubik A, Kancir J, Kosatsky T, Schwandt M. Social connection as a public health adaptation to extreme heat events. Canadian Journal of Public Health. 2020;111(6):876–9.

38. Nunes AR. The contribution of assets to adaptation to extreme temperatures among older adults. PLOS ONE. 2018;13(11):e0208121.

39. Beckmann SK, Hiete M. Predictors Associated with Health-Related Heat Risk Perception of Urban Citizens in Germany. International Journal of Environmental Research and Public Health. 2020;17(3):874.

40. Bittner M-I, Stössel U. Perceptions of heatwave risks to health: results of an qualitative interview study with older people and their carers in Freiburg, Germany. GMS Psycho-Social-Medicine. 2012;9.

41. Loughnan ME, Carroll M, Tapper N. Learning from our older people: pilot study findings on responding to heat. Australas J Ageing. 2014;33(4):271–7.

42. Valois P, Talbot D, Bouchard D, Renaud J-S, Caron M, Canuel M, et al. Using the theory of planned behavior to identify key beliefs underlying heat adaptation behaviors in elderly populations. Population and Environment. 2020;41(4):480–506.

43. House of Commons Environmental Audit Committee. Heat resilience and sustainable cooling. Fifth REport of Session 2023 - 24. London, UK; 2024.

